# VSV-EBOV vaccination induces cross-reactive T-cell responses against Bundibugyo virus in humans

**DOI:** 10.64898/2026.09.25.26363765

**Authors:** Leonie Mayer, Rolanda Londt, Eva Gruener, My Linh Ly, Marylyn M. Addo, Christof Geldmacher

## Abstract

Bundibugyo ebolavirus (BDBV) is causing an ongoing outbreak in the Democratic Republic of Congo (DRC) with no licensed vaccine available. Recent reports have shown the presence of BDBV-cross-reactive antibody responses in humans vaccinated against the related Ebola virus (EBOV). Here, we used an IFN-γ ELISpot assay to assess T cell responses against the EBOV, BDBV and SUDV glycoproteins in clinical trial participants vaccinated with the licensed VSV-EBOV vaccine. We could show that BDBV-cross-reactive T cell responses are detectable in the majority of individuals and that conserved regions within the glycoprotein are targeted. In the absence of a BDBV-specific vaccine, these data support efficacy trials of the VSV-EBOV vaccine during the current outbreak.

---

The ongoing 2026 outbreak of Bundibugyo ebolavirus (BDBV) in the Democratic Republic of the Congo (DRC) is currently the second largest Orthoebolavirus outbreak globally with confirmed cases exceeding 6,000 and a case fatality rate of 48% and no licensed vaccine available.^1^ rVSVΔG-ZEBOV-GP (Ervebo; Merck Sharp & Dohme, here abbreviated as VSV-EBOV) is a highly efficacious vaccine against the phylogenetically related Ebola virus (EBOV, formerly Zaire ebolavirus), consisting of a recombinant Vesicular Stomatitis Virus (VSV) viral vector that encodes the glycoprotein (GP) of the Kikwit 1995 EBOV isolate^2^. Immunization with VSV-EBOV conferred partial cross-protection against lethal, intramuscular BDBV challenge in non-human primates (NHPs)^3^ and recent studies could show detectable, BDBV-reactive antibody responses in VSV-EBOV-vaccinated individuals^4–6^. Although vaccine-induced, EBOV-GP-binding antibodies are considered a putative correlate of protection against EBOV disease^7^, it remains unknown, if these BDBV-cross-reactive responses are sufficient to protect humans against BDBV disease^8^.

Virus-specific T cell responses associate with virus control and protection against severe diseases, including EBOV disease^9^, and can provide broader cross-reactivity and greater resilience to viral escape by targeting multiple, often conserved, linear peptide epitopes^10–12^. Whether VSV-EBOV vaccination induces BDBV-reactive T cells in humans is, however, not known. To address the extent of pan-Orthoebolavirus-specific T cell recognition upon vaccination, we analyzed and mapped T cell reactivity against EBOV, BDBV and SUDV in VSV-EBOV recipients (ClinicalTrials.gov; NCT02283099, **Fig. 1a**)^13,14^ and further investigated whether conserved regions within the GP protein are targeted.

**Figure 1.**
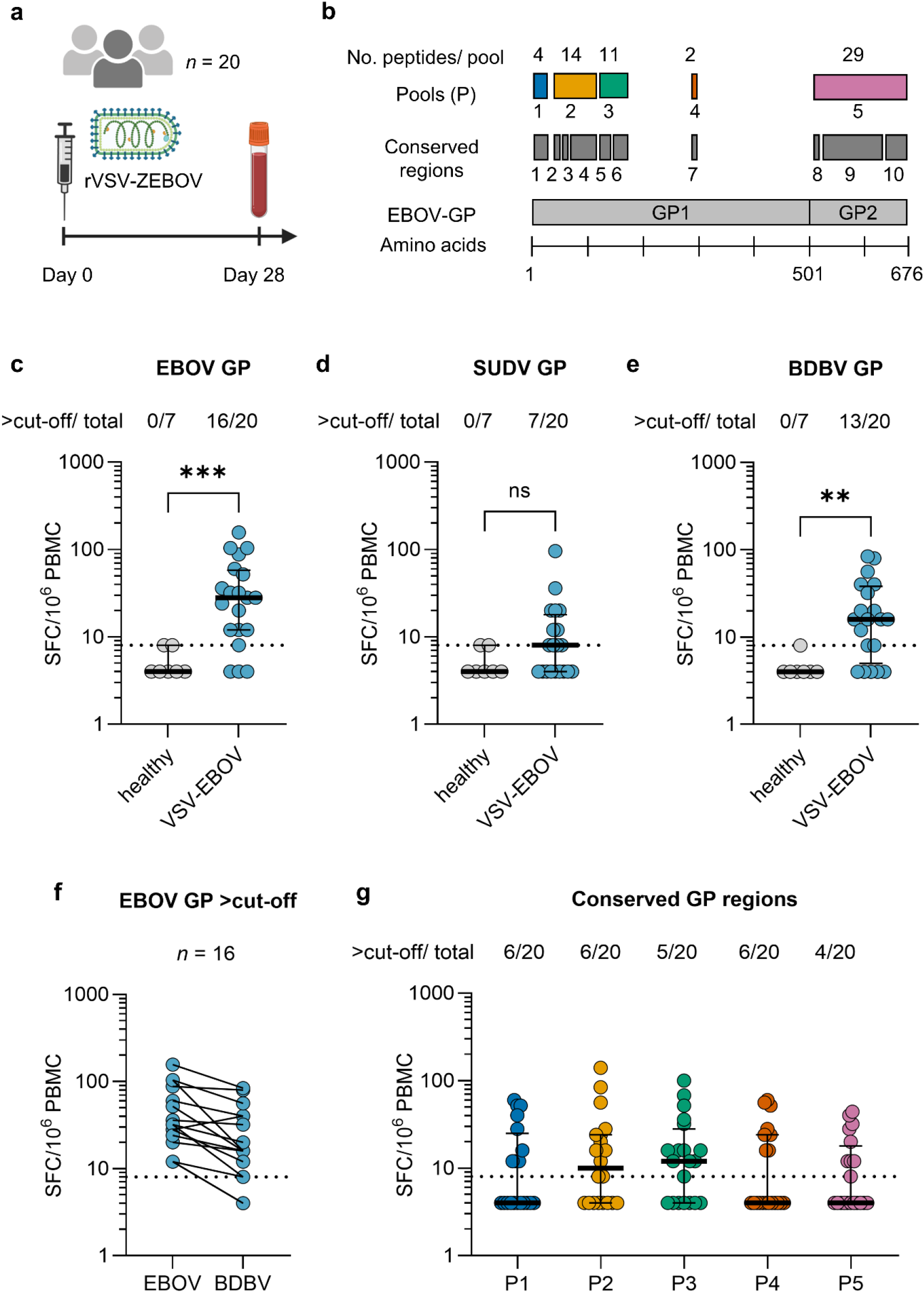
Orthoebolavirus-cross-reactive T cell responses following VSV-EBOV vaccination. a) Study design: Healthy adults received a single intramuscular dose of the rVSVΔG-ZEBOV-GP vaccine (VSV-EBOV) on day 0 and blood samples were collected on day 28 to analyze T cell responses (Created in BioRender. Mayer, L. (2026) https://BioRender.com/6dmmwia). b) Peptide pools: Schematic of the EBOV GP sequence showing EBOV/ SUDV/ BDBV conserved regions 1-7 and 8-10 within the GP1 and GP2, respectively. Pools 1-5 consisting of overlapping peptides covering these conserved regions were generated (see supplementary appendix for pooling strategy). T cell response measured by IFN-γ ELISpot in healthy controls (n = 7) and VSV-EBOV-vaccinated individuals (n = 20) against the EBOV-GP (c), SUDV-GP (d), and BDBV-GP (e). The horizontal line indicates the lower limit of detection (= 8 SFC/10^6^ PBMCs). Number of individuals with a positive response, defined as being above the stimulus-specific cut-off of the mean + 2* standard deviation healthy control response, is indicated above. f) Comparison of EBOV- and BDBV-GP-specific T cell responses in individuals with a positive EBOV response (n = 16). g) T cell response of VSV-EBOV-vaccinated individuals against conserved pools 1-5, with number of individuals above the cut-off. SFC/ 10^6^ PBMCs = spot-forming cells per million peripheral blood mononuclear cells. ns = not significant, *** p < 0.001, ** p < 0.01.

The EBOV and BDBV GPs are phylogenetically distant (66.3 % amino acid identity), but more similar than the EBOV and SUDV GPs (56.3 % amino acid identity (**Fig. S1)**. By multiple sequence alignment (see appendix for method details) we identified several amino acid sequences that are highly conserved between the GPs of EBOV, SUDV and BDBV and generated peptide pools covering these regions (**Fig. 1b**). We then assessed cross-reactive T cell responses at day 28 post VSV-EBOV vaccination in 20 vaccine recipients by IFN-γ ELISpot (see appendix for method details). Seven healthy controls were included for comparison. With a median frequency of 28 SFC/10^6^ PBMC, 16 out of 20 vaccine recipients showed EBOV-GP-specific T cell responses 28 days post vaccination (**Fig. 1c**). While only 7 out of 20 individuals responded to the SUDV GP (median of 8 SFC/10^6^ PBMC, **Fig. 1d**), 13 out of 20 individuals had BDBV-GP-reactive T cell responses (median 16 SFC/10^6^ PBMC, **Fig. 1e**). T cell responses were significantly higher in vaccinees compared to healthy controls against EBOV-GP (p = 0.0009) and BDBV-GP (p = 0.0041), but not against SUDV GP (p = 0.15). Most vaccinees with a positive EBOV-GP response also had BDBV-reactive T cells (**Fig. 1f**) with positively correlating T cell frequencies (Spearman r = 0.77, p = 0.0007, **Fig. S2**). Recognition of five peptide pools representing all conserved GP regions were analyzed in parallel, revealing that these regions are recognized by 4-6 of the 20 analyzed vaccine recipients (**Fig. 1g**).

Our study shows that a single immunization with the available, licensed EBOV vaccine induces considerable T cell cross-reactivity against BDBV and to a lesser extent against SUDV. While it is not known, whether these T cells contribute to improved clinical outcomes upon BDBV exposure in humans, preclinical studies show that T cell responses are required for vaccine-mediated protection in both NHP and rodent models^15–17^. CD4 T cell help to B cells is required for vaccine-induced protection against EBOV disease of NHPs^18^, while depletion and adoptive cell transfer studies have demonstrated a protective role of CD8 T cells in rodents^15,16^.

Although limited to few participants, our findings provide first evidence that VSV-EBOV vaccination against EBOV induces BDBV-cross-reactive T cell responses. In the absence of a BDBV-specific vaccine, these data support the systematic evaluation of the available EBOV vaccine within clinical trials during the current outbreak.

## Data Availability

Individual participant data that underlie the results reported in this article, after de-identification (text, tables, figures, and appendices), will be available immediately following publication to researchers who provide a methodologically sound proposal to achieve aims specified in the approved proposal. Proposals should be directed to Leonie Mayer. To gain access, data requestors will need to sign a data access agreement.

## Contributors

Conceptualization – LM, CG, MMA. Methodology – LM, MLL, EG, RL. Formal analysis – LM, MLL, RL, EG. Investigation - LM, MLL, RL, EG. Resources – CG, MMA. Writing – original draft – LM, CG. Writing – review & editing – LM, RL, EG, MLL, CG, MMA. Visualization – LM, RL. Supervision – CG, MMA. All authors had full access to the data and final responsibility for submission. LM and CG have accessed and verified the underlying data. All authors reviewed and approved the final version of the manuscript.

## Data sharing

Individual participant data that underlie the results reported in this article, after de-identification (text, tables, figures, and appendices), will be available immediately following publication to researchers who provide a methodologically sound proposal to achieve aims specified in the approved proposal. Proposals should be directed to the corresponding author. To gain access, data requestors will need to sign a data access agreement.

## Acknowledgements

This study was supported by the EUROPEAN HEALTH AND DIGITAL EXECUTIVE AGENCY (Grant number 101202831 — EVH — EU4H-2024-PJ-01: European Vaccine Hub for pandemic preparedness), the German Center for Infection Research (DZIF) and the Deutsche Forschungsgemeinschaft (DFG, German Research Foundation) –SFB 1648/1 2024 – 512741711.

## Supplementary appendix

## Methods

### Peptide Design and pooling

Pools of 15mer peptides overlapping by 10 amino acids were designed and synthesized by Peptides & Elephants GmbH (Henningsdorf, Germany) for clinical isolates representing the 2026 BDBV strain collected in the Democratic Republic of the Congo (Pathoplexus accession PP_006XXY5.1), the 2025 EBOV strain collected in Bulape (PP_004BPQ4.1), and the 2025 SUDV strain collected in Uganda (PP_0011CQ4.2).

### Design of peptide pools for conserved regions

Conserved regions were identified through multiple sequence alignment of the full-length Glycoprotein sequences from the clinical isolates used for peptide design (Supplementary Figure 1). Alignments were generated with Clustal Omega v1.2.4 (default parameters) and analyzed in Jalview v2.11.5.2. Conservation at each alignment position was quantified using Jalview’s default conservation annotation based on the AMAS method of Livingstone and Barton, yielding a physicochemical property conservation index ranging from 0 to 11. Amino acid positions with a conservation index ≥ 8 were considered highly conserved. Conserved regions were defined by eight or more consecutive amino acids with a conservation index of ≥ 8. Based on this algorithm, 10 conserved regions along the Glycoprotein were defined.

Using the BDBV 2026 sequence, 15-mers spanning the whole GP protein were designed. Individual peptides containing at least eight amino acids from a conserved region were selected for inclusion in the pools. Peptides were grouped based on their proximity, with no more than 30 peptides per pool. Five conserved region (CR) pools were created: CR pools 1 - 3 covered conserved regions located within the N-terminal region of GP1, CR pool 4 covered a central region of GP1 and CR pool 5 combined conserved regions in GP2.

### Study samples

Twenty participants of a previously published open-label, dose-escalation phase 1 clinical trial of the rVSVΔG-ZEBOV-GP vaccine (ClinicalTrials.gov; NCT02283099)^1,2^ and seven healthy controls (approved by the Ethics Committee of the Hamburg Medical Association, reference number: PV4780) were included in this study. All trial participants and healthy controls provided written informed consent. Trial participants had received a single intramuscular vaccination of either 3 × 10^5^ plaque-forming units (PFU, n = 9), 3 × 10^6^ PFU (n = 4) or 2 × 10^7^ PFU (n = 8) of the vaccine. Peripheral blood mononuclear cells (PBMCs) were isolated by ficoll density gradient centrifugation from whole blood collected at day 28 post vaccination and cryopreserved.

### Measurement of Orthoebolavirus-specific T cell responses by IFN-γ ELISpot

T cell responses were analyzed by IFN-γ ELISpot. After overnight resting, 1.25 × 10^5^ PBMCs/ well were incubated with overlapping peptide pools (2 μg peptide/ml in 0.25-0.5% dimethyl sulfoxide (DMSO)), Phytohemagglutinin (positive control) or a cytomegalovirus–Epstein-Barr–influenza virus pool (JPT, positive control) or culture medium with 0.25-0.5% DMSO (negative control) 16 h at 37°C and 5% CO_2_. Spot-forming cells (SFC) per million PBMCs were counted using an AID ELISpot Reader System. The results are reported as the background (DMSO)–subtracted mean count of duplicate wells.

### Data analysis

Statistical analysis, data summary and visualization were done in Microsoft Excel (v1808), GraphPad Prism (v10.4.2) and BioRender.com. Statistical tests were two-sided and a p-value of ≤0.05 was considered statistically significant. Groups were compared by Mann-Whitney test. Correlation analysis was done using Spearman r.

## Supplementary Figures

**Figure S1.**
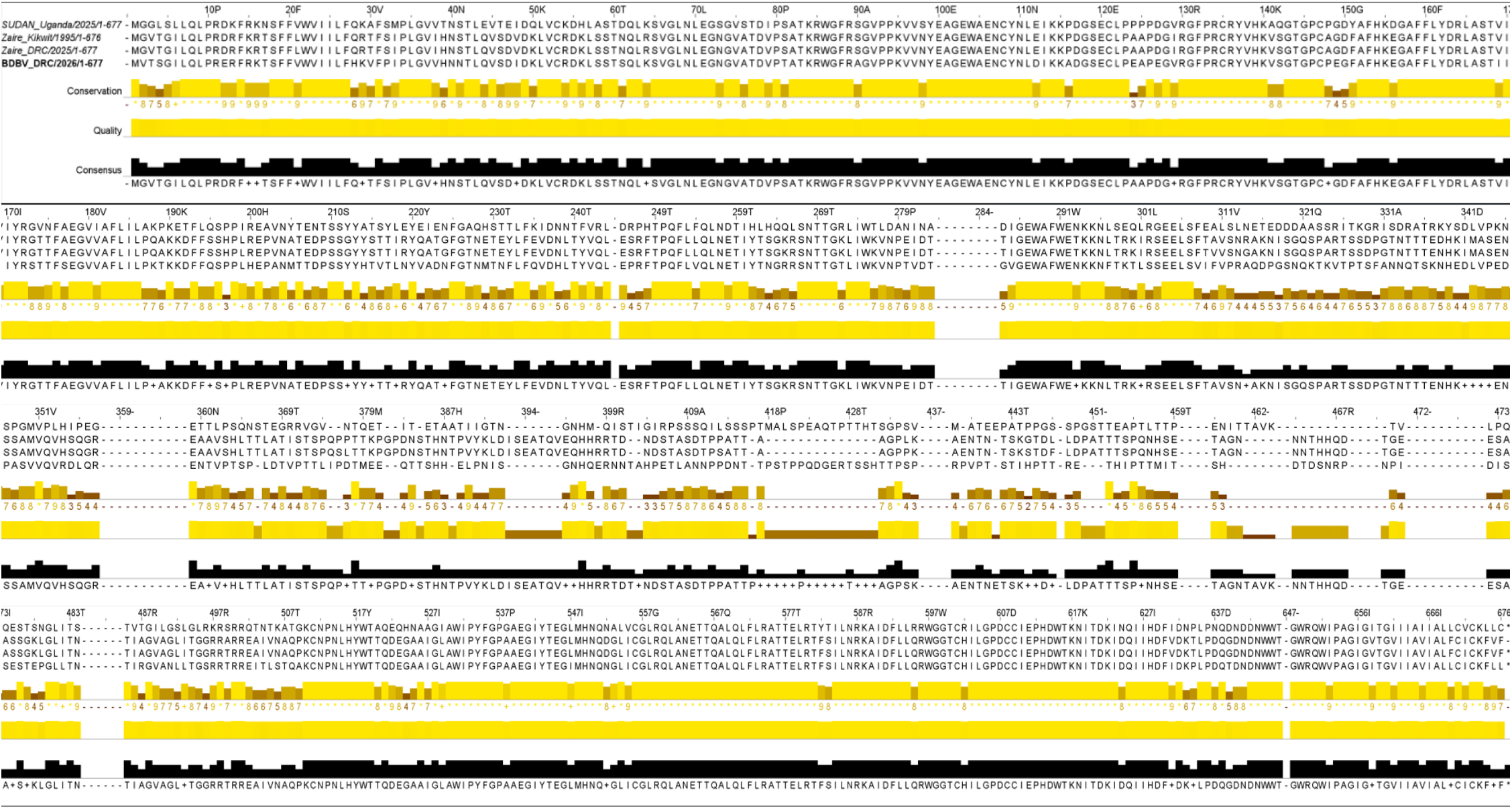
Multiple-sequence alignment and conservation analysis of glycoprotein sequences from the clinical BDBV, EBOV, and SUDV isolates used for peptide design.

**Figure S2.**
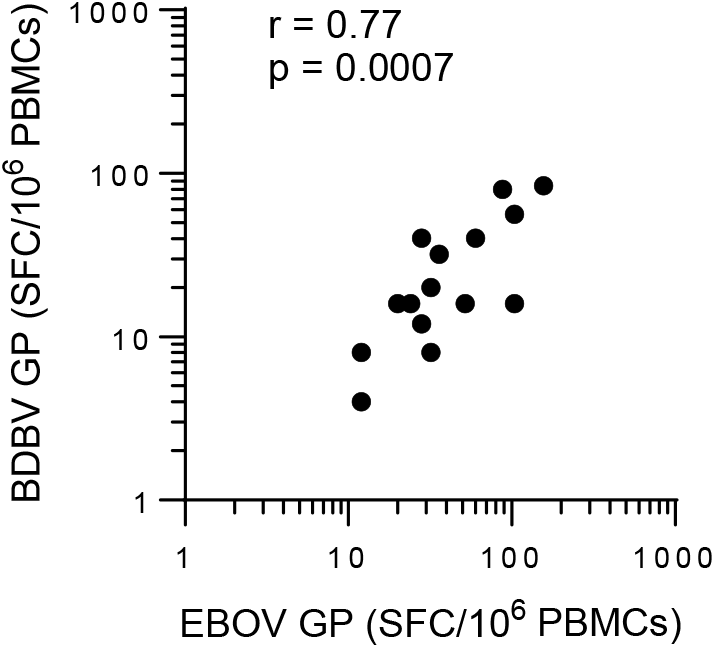
Correlation of EBOV-GP- and BDBV-GP-specific T cell frequencies in vaccinees who showed a positive EBOV GP response (n = 16). Correlation was assessed by Spearman r. SFC/ 10^6^ PBMCs = spot-forming cells per million peripheral blood mononuclear cells.

